# Large-Scale Deep Learning for Metastasis Detection in Pathology Reports

**DOI:** 10.1101/2024.12.12.24318789

**Authors:** Patrycja Krawczuk, Zachary R Fox, Valentina Petkov, Serban Negoita, Jennifer Doherty, Antoinette Stroupe, Stephen Schwartz, Lynne Penberthy, Elizabeth Hsu, John Gounley, Heidi A. Hanson

**Author notes:** Equal contribution (Patrycja Krawczuk, Zachary R Fox).

## Abstract

No existing algorithm can reliably identify metastasis from pathology reports across multiple cancer types and the entire US population. In this study, we develop a deep learning model that automatically detects patients with metastatic cancer by using pathology reports from many laboratories and of multiple cancer types. We trained and validated our model on a cohort of 29,632 patients from four Surveillance, Epidemiology, and End Results (SEER) registries linked to 60,471 unstructured pathology reports. Our deep learning architecture trained on task-specific data outperforms a general-purpose LLM, with a recall of 0.894 compared to 0.824. We quantified model uncertainty and used it to defer reports for human review. We found that retaining 72.9% of reports increased recall from 0.894 to 0.969. This approach could streamline population-based cancer surveillance to help address the unmet need to capture recurrence or progression.

## Introduction

Survival rates after an initial cancer diagnosis have improved drastically over the past 50 years. There are currently an estimated 18.1 million cancer survivors in the United States, and this number is projected to rise to 22.5 million by 2032^1^. Despite these advances, US population-based cancer surveillance is focused primarily on cancer mortality, thereby hindering our ability to assess long-term survivor outcomes. Metastasis, a critical indicator of disease progression and recurrence, remains difficult to monitor at a population scale because there are no explicit federal mandates for collecting this information, and the required extended follow-up period strains already limited hospital and registry resources.

The National Cancer Institute’s (NCI) Surveillance, Epidemiology, and End Results (SEER) program has a robust infrastructure for collecting and utilizing pathology reports for cancer incidence ascertainment. In this paper, we demonstrate how this infrastructure can be leveraged to expand the collection of information about metastatic disease, thus addressing an unmet need by cancer patients, clinicians, researchers, and policymakers. By expanding the scope of data collection, this work will enhance our understanding of metastatic disease and ultimately inform cancer treatment strategies and survivorship care. To this end, the rapid and accurate summarization of large volumes of unstructured information is crucial for effective population-level cancer surveillance. Natural language processing (NLP) facilitates the real-time analysis and automated classification of clinical texts^2–6^. NLP approaches typically require numerous pathology reports paired with labels to train models that accomplish these objectives. However, labeled data is scarce in the medical domain and studies that leverage NLP to identify metastases from pathology reports typically focus on a single cancer type^12–16^.

General-purpose large language models (LLMs) enable a new paradigm for automated summarization of unstructured data. Unlike traditional NLP approaches, LLMs are pre-trained for general use and can be employed without labeled data to extract information across a broad set of tasks using carefully crafted prompts, a technique known as zero-shot learning. A recent study evaluated the use of LLMs in the zero-shot setting to classify breast cancer pathology reports, demonstrating superior performance over task-specific supervised models^12^ with limited annotated data. This result demonstrates the advantage of LLMs for tasks with limited annotated data. Although LLMs hold great promise^13–15^, methods for validating their predictions are still scarce^16,17^, and their use for text classification remains largely unexplored^18^.

The reliability of predictions from NLP algorithms, including those used in LLMs, can vary significantly, with some predictions being confident and others uncertain. This uncertainty is often overlooked due to the predominant focus on the overall accuracy of model results rather than a nuanced understanding of individual predictions. For confident predictions, automated classification can aid in population surveillance by enabling low-cost information extraction. For unconfident predictions, case identification can be facilitated through human-machine collaboration for unconfident predictions.

This study compares a task-specific deep learning approach with a general LLM for automatically detecting a metastatic disease diagnosis for five common cancer types in unstructured clinical pathology reports from local and national laboratories (Figure 1: Data Gathering). Using these 60,471 pathology reports from 29,632 patients, we sought to understand (1) if a task-specific deep learning model trained on the annotated reports can accurately identify metastatic disease in a population-scale dataset, (2) how uncertainty in its predictions can be evaluated and appropriately managed, and (3) if this task-specific model can outperform the zero-shot performance of a pre-trained LLM. In contrast to previous studies, the patient data came from numerous laboratories/hospital systems across multiple regions in the United States, leading to wide variability in the format, patient pools, and pathologists who drafted these reports.

**Figure 1:**
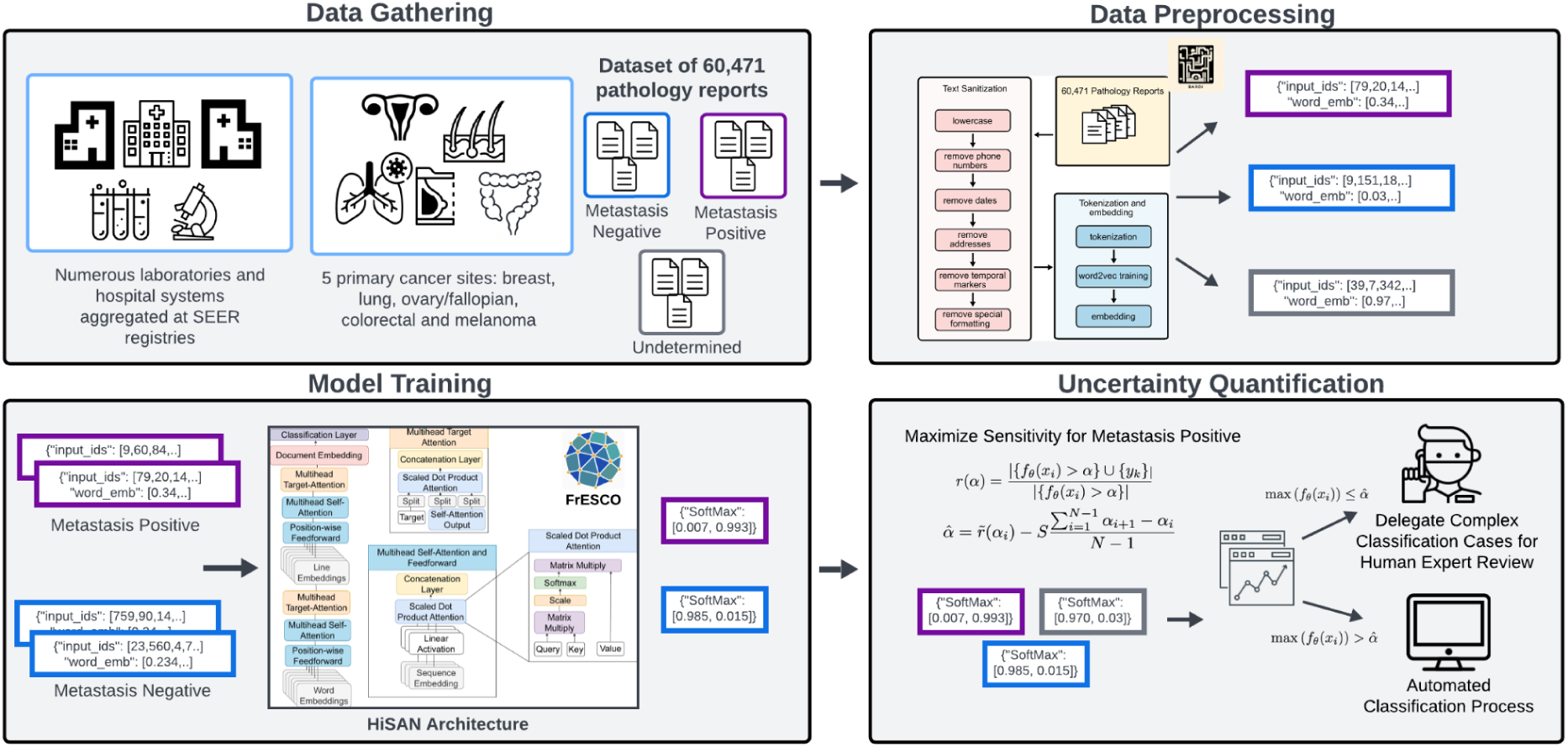
The proposed framework processes pathology reports aggregated from numerous laboratories and hospital systems within SEER registries, focusing on five primary cancer sites: breast, lung, ovary/fallopian, colorectal, and melanoma (**Data Gathering**). Oncology data experts annotate each report as Metastasis Positive, Metastasis Negative, or Undetermined, creating a labeled dataset for model training. During **Data Preprocessing**, the raw text data is converted into tokenized embeddings using BARDI. A deep learning model based on the HiSAN architecture is then trained to classify reports as either Metastasis Positive or Metastasis Negative (**Model Training**). The model utilizes multi-headed self-attention and generates class probabilities. In the **Uncertainty Quantification** stage, softmax probabilities are evaluated to set a sensitivity-maximizing threshold for Metastasis Positive cases. Patient-level uncertainty is quantified by comparing the model’s confidence scores against this threshold. Cases with certainty scores below the threshold are flagged for expert review to ensure accuracy, while cases above the threshold are classified automatically.

## Results

First, we present the characteristics of the dataset used in this study. Second, we present the results of training the task-specific HiSAN model and evaluate its performance across different confidence thresholds by utilizing 5-fold cross-validation (see Table S1). This evaluation allows us to select a threshold that balances the model’s performance with the number of reports marked for a second review. After selecting the optimal threshold, we compare our task-specific HiSAN model with the general-purpose Llama3 across different cancer types. Finally, we analyze the task-specific model benefits from training on multiple cancer types by comparing it to a set of models trained on individual cancer types.

### Dataset

The dataset consists of 60,471 individual pathology reports from 29,632 patients. The reports were processed at four SEER registries in Utah, New Jersey, Seattle/Puget Sound, and Louisiana between 2019 and 2023. The reports were selected from those undergoing routine cancer reportability screening at each registry over a 1-year period (from February 2022 to February 2023). The following criteria were used: First, the pathology reports had to be linked to patients with cancer in one or more of five primary sites (breast, lung, ovary, colorectal, and/or melanoma). Second, the report must have had a specimen collection date of more than 120 days after the diagnosis. Some reports were reclassified during a second review, resulting in 551 reports associated with *Other* cancers. To create gold<u>-</u>standard labels, the oncology data specialists annotated each report as *metastasis negative*, *metastases positive*, or *metastasis undetermined*. The dataset was preprocessed as described in the Supplementary Material. Cancer registries obtain race from various sources, including medical records, pathology reports, and administrative databases.

Table 1 presents a detailed breakdown of the pathology report dataset categorized by metastasis status across various demographic and clinical groups. Most reports in this dataset are from the Seattle registry, accounting for 26,507 out of 60,471 total reports, and provide information about nearly 13,000 unique patients. Among racial groups, 27.4% of the reports for White patients and 31.4% of the reports for Black patients are associated with metastasis. The proportion of metastatic reports decreases with age: 32.1% of reports for patients under 44 are *metastasis positive* compared to 24.9% for patients between 45 and 70 years old and 20.8% for 71 and older. Most of the reports are from patients aged 45–70, which is consistent with general cancer trends in the United States^20^. Over 70% of the cases involve female patients, with the dataset predominantly featuring breast malignancies. Most of these breast cancer cases are non-metastatic, driving an imbalance of classes.

**Table 1:** Number of pathology reports with given metastasis status across various demographic and clinical groups.

| No. (%) | *Number of Patients*<br>(N=29,632) | *Total Pathology Reports*<br>(N= 60,471) | *Metastasis Negative*<br>(n=43,286<br>[71.58%]) | *Metastasis Positive*<br>(n=14,959<br>[24.74%]) | *Metastasis Undetermined*<br>(n=2,226<br>[3.68%]) |
| --- | --- | --- | --- | --- | --- |
| **All Registries** |  |  |  |  |  |
| Seattle | 12,732 (42.97) | 26,507 (43.83) | 21,277 (49.15) | 4,800 (32.09) | 430 (19.32) |
| New Jersey | 8,607 (29.05) | 16,770 (27.73) | 10,833 (25.03) | 5,163 (34.51) | 774 (34.77) |
| Louisiana | 5,723 (19.31) | 11,886 (19.66) | 7,689 (17.76) | 3,451 (23.07) | 746 (33.51) |
| Utah | 2,570 (8.67) | 5,308 (8.78) | 3,487 (8.06) | 1,545 (10.33) | 276 (12.40) |
| **Sex** |  |  |  |  |  |
| Female | 20,765 (70.08) | 43,102 (71.28) | 30,634 (70.77) | 10,896 (72.84) | 1,572 (70.62) |
| Male | 8,858 (29.89) | 17,359 (28.71) | 12,644 (29.21) | 4,061 (27.15) | 654 (29.38) |
| **Age at Diagnosis** |  |  |  |  |  |
| Under 44 | 3,890 (13.13) | 7,817 (12.92) | 5,305 (12.26) | 2,303 (15.40) | 209 (9.39) |
| Ages 45 to 70 | 19,949 (67.32) | 41,015 (67.83) | 29,232 (67.53) | 10,227 (68.37) | 1,556 (69.90) |
| 71 or Older | 5,793 (19.55) | 11,639 (19.25) | 8,749 (20.21) | 2,429 (16.24) | 461 (20.71) |
| **Race** |  |  |  |  |  |
| White | 24,310 (82.04) | 49,991 (82.67) | 36,274 (83.80) | 11,950 (79.89) | 1,767 (79.38) |
| Black | 3,035 (10.24) | 6,186 (10.23) | 3,880 (8.96) | 1,941 (12.98) | 365 (16.40) |
| Other/Unknown | 2,287 (7.72) | 4,294 (7.10) | 3,132 (7.23) | 1068 (7.14) | 94 (4.22) |
| **Cancer Type** |  |  |  |  |  |
| Breast | 13,227 (44.64) | 27,607 (45.65) | 19,769 (45.67) | 6,918 (46.25) | 920 (41.33) |
| Melanoma | 5,866 (19.80) | 11,710 (19.36) | 9,949 (22.98) | 1,480 (9.89) | 281 (12.62) |
| Lung | 4,034 (13.61) | 8,713 (14.41) | 5,651 (13.06) | 2,493 (16.67) | 569 (25.56) |
| Colorectal | 5,517 (18.62) | 10,513 (17.39) | 6,825 (15.78) | 3,353 (22.41) | 335 (15.05) |
| Ovarian/Fallopian | 706 (2.38) | 1,377 (2.28) | 679 (1.57) | 604 (4.04) | 94 (4.22) |
| Other | 282 (0.95) | 551 (0.91) | 413 (0.95) | 111 (0.74) | 27 (1.21) |

### Task-Specific HiSAN Approach

We present the results of the three-class classification without applying confidence thresholds for abstention in Table 2 and Fig. 3B. The model achieves a mean precision of 0.864 and recall of 0.894. These results are negatively impacted by the presence of the *undetermined* class in the test set that the model is forced to classify as either *metastasis positive* or *metastasis negative.* For the results that consist only of *metastasis positive* and *metastasis negative*, see Table S2 in Supplementary Materials.

**Table 2:** Comparison of a HiSAN model trained on all cancers vs. HiSAN models trained on individual cancers.

|  | *HiSAN Evaluation on Three Classes* |  |  | *HiSAN Trained on each Primary Site Separately Evaluation on Three Classes* |  |  |
| --- | --- | --- | --- | --- | --- | --- |
|  | *Precision*<br>*Mean*<br>*(Low, High)* | *Recall*<br>*Mean*<br>*(Low, High)* | *F1*<br>*Mean*<br>*(Low, High)* | *Precision*<br>*Mean*<br>*(Low, High)* | *Recall*<br>*Mean*<br>*(Low, High)* | *F1*<br>*Mean*<br>*(Low, High)* |
| *All Data* | 0.864<br>(0.856,0.868) | 0.894<br>(0.890,0.899) | 0.878<br>(0.872,0.883) | NA | NA | NA |
| Breast | 0.879<br>(0.872,0.884) | 0.905<br>(0.899,0.909) | 0.890<br>(0.886,0.894) | 0.876<br>(0.871,0.881) | 0.904<br>(0.901,0.909) | 0.889<br>(0.885,0.893) |
| Melanoma | 0.902<br>(0.886,0.923) | 0.920<br>(0.909,0.936) | 0.910<br>(0.897,0.929) | 0.890<br>(0.877,0.900) | 0.909<br>(0.903,0.915) | 0.899<br>(0.889,0.903) |
| Lung | 0.785<br>(0.762,0.807) | 0.841<br>(0.827,0.862) | 0.812<br>(0.797,0.834) | 0.782<br>(0.758,0.805) | 0.838<br>(0.819,0.851) | 0.809<br>(0.791,0.825) |
| Colorectal | 0.876<br>(0.849,0.885) | 0.902<br>(0.877, 0.912) | 0.888<br>(0.862,0.898) | 0.868<br>(0.831,0.880) | 0.895<br>(0.860,0.909) | 0.881<br>(0.845,0.894) |
| Ovarian/<br>Fallopian | 0.709<br>(0.613,0.835) | 0.763<br>(0.674,0.875) | 0.733<br>(0.641,0.854) | 0.577<br>(0.516,0.610) | 0.581<br>(0.549,0.635) | 0.550<br>(0.525,0.619) |
| Other | 0.823<br>(0.631, 0.913) | 0.862<br>(0.744,0.930) | 0.839<br>(0.682,0.917) | 0.644<br>(0.431,0.816) | 0.734<br>(0.513,0.806) | 0.678<br>(0.442,0.810) |

### Confidence Threshold Determination for Task-Specific HiSAN Approach

Next, we evaluate the model at different confidence thresholds to determine how precision and recall are related. We score the model at threshold values ranging from 0.5 to 1.0. Any prediction with the highest softmax score below the threshold is excluded from the performance metrics.

We refer to the proportion of these omitted reports as the *abstained percentage*. Figure 2 (top row) illustrates the relationship between the precision and recall for the *metastasis negative* (Fig. 1A) and *metastasis positive* (Fig. 2B) classes at each threshold. The error bars denote one standard deviation from the mean. In the case of *metastasis negative*, the precision and recall are close to the unity line, indicating a relatively equal number of false positives and false negatives. The recall for the *metastasis positive* class is always higher than the precision, indicating fewer false negatives than false positives. Figure 2 (bottom row) shows the recall (C) and precision (D) as a function of the abstained percentage at a given confidence threshold for the model. All precision and recall curves show a steep increase with a small loss of reports, followed by a plateau at which further increases in precision or recall require a significant reduction in the number of retained reports. All curves are monotonically increasing, demonstrating that the model is correct about reports in which it is most confident.

**Figure 2:**
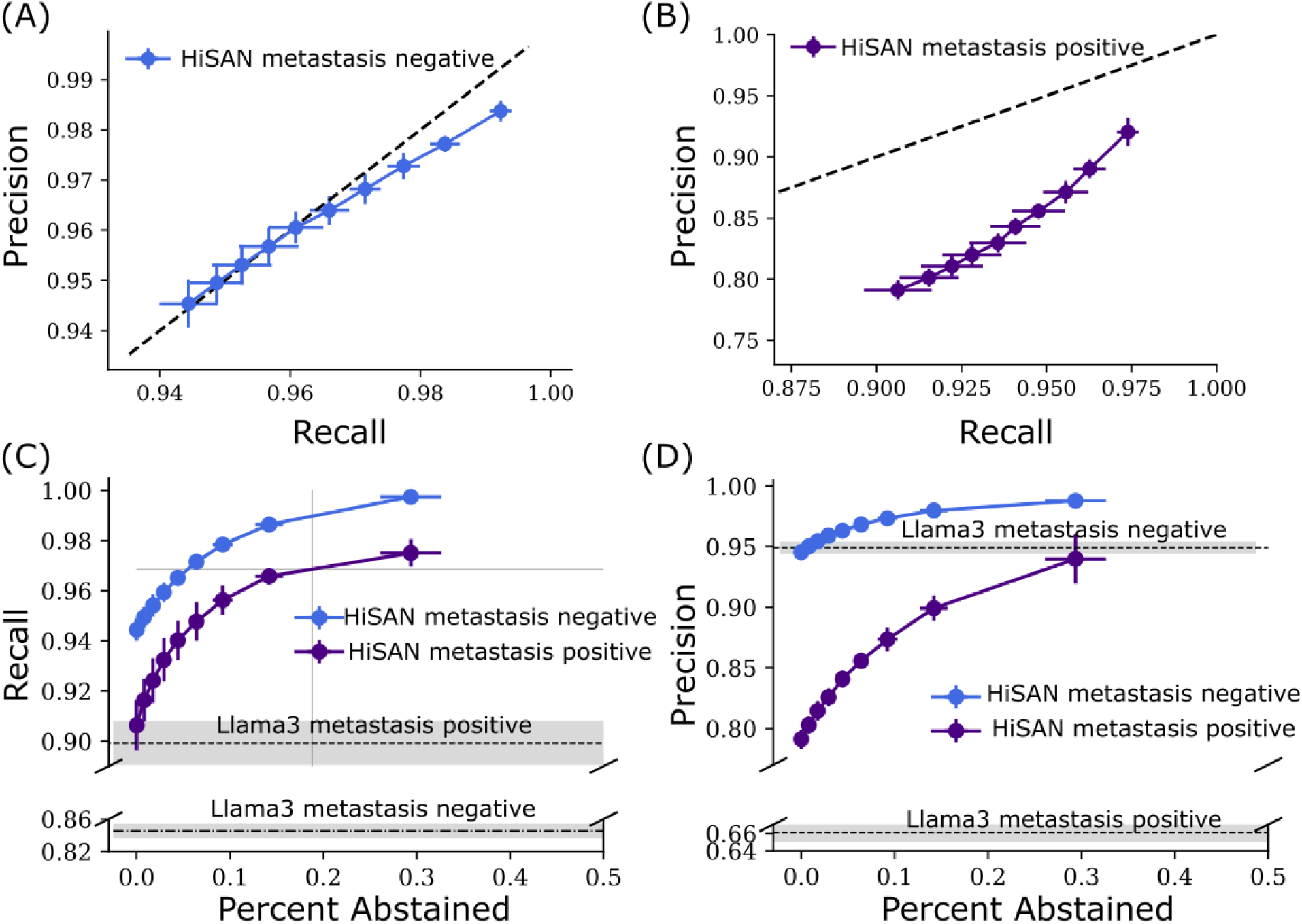
HiSAN precision-recall-abstention curves for metastasis negative (A) and metastasis positive (B). Lines indicate validations for each of the experiments at different abstention thresholds. Each dot indicates the average across the five validation results, and the error bars indicate ±1 standard deviation. The dashed-black lines are the unity line and indicate equal precision and recall. Recall (C) and precision (D) on validation data for HiSAN and Llama3. HiSAN results are shown as a function of the fraction of data removed to achieve a given recall/precision. Each dot indicates the average across the five validation results, and the error bars indicate ±1 standard deviation. For Llama3 (which has no abstention mechanism), the horizontal-dashed lines and shaded region indicate the average recall/precision ±1 standard deviation.

We define the optimal confidence threshold as the knee point in the nonlinear recall-abstention curve for the *metastasis positive* class (i.e. the blue line in Fig. 2C). To this end, we use a knee-finding algorithm^21^ (see Methods), identifying a threshold of 0.955. At this threshold, the model achieves a mean precision of 0.951 (see Table 3) and mean recall of 0.969 with a mean abstention rate of 0.271.

**Table 3:** Comparison of a HiSAN model with abstention vs. Llama3.

|  | **HiSAN Three Classes with Threshold of 0.955** |  |  | **Llama3 Zero-Shot Learning on Three Classes** |  |  |
| --- | --- | --- | --- | --- | --- | --- |
|  | **Precision**<br>Mean<br>(Low, High) | **Recall**<br>Mean<br>(Low, High) | **F1**<br>Mean<br>(Low, High) | **Precision**<br>Mean<br>(Low, High) | **Recall**<br>Mean<br>(Low, High) | **F1**<br>Mean<br>(Low, High) |
| **All Data** | 0.951<br>(0.941,0.960) | 0.969<br>(0.964,0.974) | 0.960<br>(0.952,0.967) | 0.842<br>(0.833,0.851) | 0.824<br>(0.817,0.832) | 0.825<br>(0.816,0.834) |
| Breast | 0.956<br>(0.942,0.965) | 0.973<br>(0.963,0.978) | 0.964<br>(0.952,0.971) | 0.852<br>(0.843,0.861) | 0.836<br>(0.828,0.845) | 0.837<br>(0.829,0.846) |
| Melanoma | 0.962<br>(0.950,0.973) | 0.976<br>(0.967,0.982) | 0.968<br>(0.959,0.977) | 0.896<br>(0.885,0.907) | 0.850<br>(0.833,0.867) | 0.863<br>(0.848,0.877) |
| Lung | 0.918<br>(0.900,0.944) | 0.950<br>(0.939,0.962) | 0.934<br>(0.919,0.953) | 0.770<br>(0.746,0.794) | 0.755<br>(0.731,0.779) | 0.749<br>(0.722,0.777) |
| Colorectal | 0.950<br>(0.940,0.957) | 0.966<br>(0.962,0.970) | 0.958<br>(0.951,0.963) | 0.850<br>(0.828,0.872) | 0.843<br>(0.825,0.861) | 0.843<br>(0.823,0.863) |
| Ovarian/<br>Fallopian | 0.919<br>(0.875,0.977) | 0.947<br>(0.919,0.979) | 0.932<br>(0.898,0.975) | 0.702<br>(0.618,0.785) | 0.712<br>(0.637,0.787) | 0.694<br>(0.613,0.774) |
| Other | 0.996<br>(0.979,1.000) | 0.996<br>(0.979,1.000) | 0.995<br>(0.977,1.000) | 0.823<br>(0.696,0.950) | 0.737<br>(0.624,0.851) | 0.758<br>(0.655,0.861) |
| Abstention Rate: | Mean: 0.271, Min: 0.232, Max: 0.320 |  |  |  |  |  |

### General LLM Approach

Next, we demonstrate our prompt-based, zero-shot classification results. Llama3 is provided Prompt 1 (see Methods) and a pathology report to classify into one of the three original classes (Fig. 3A). This method leads to an F1 score of 0.825, mean precision of 0.842 and recall of 0.824 (Table 3).

**Figure 3:**
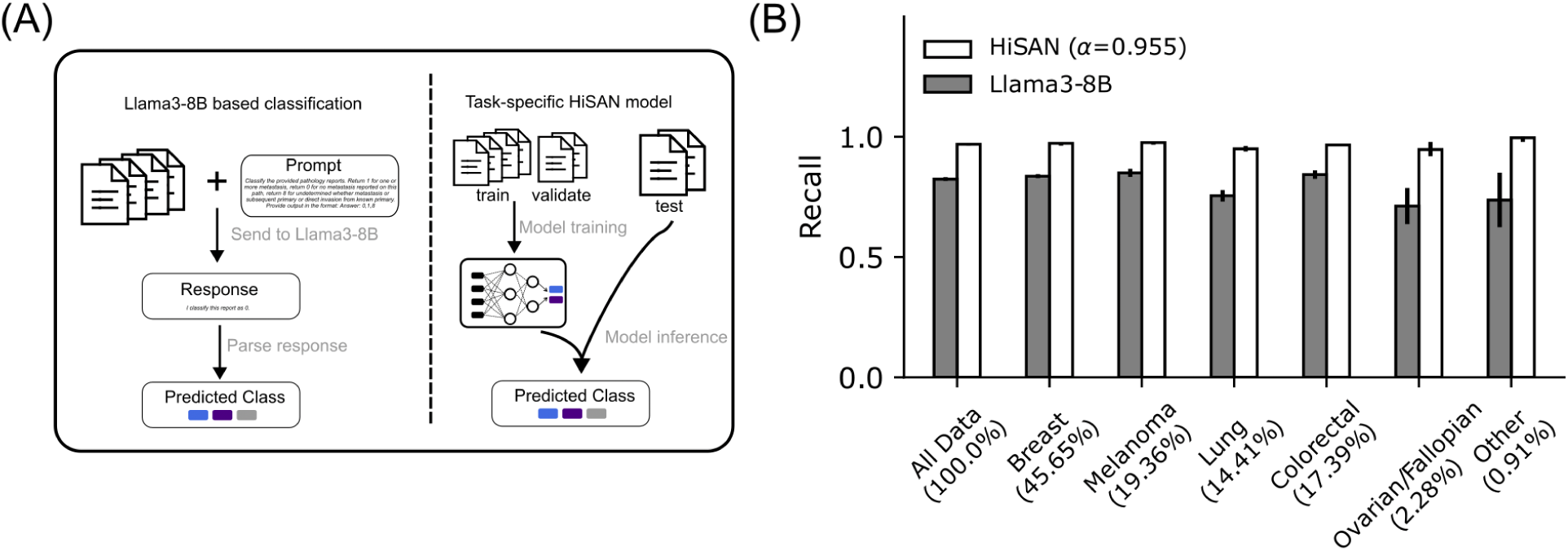
Comparing Llama3 and HiSAN for metastasis prediction. (A) Left: Workflow for zero-shot Llama3-based classification of clinical pathology reports. The pathology report along with a prompt are passed to the LLM, which returns a partially formatted response. The response is parsed and the predicted class (metastasis positive, metastasis negative, undetermined) is extracted. Right: Workflow of HiSAN-based model predictions. The model is trained and validated using 5-fold cross validation. After training, the model is evaluated on the held-out pathology reports. The reports above the determined confidence threshold α are included for evaluation. (B) Mean Recall of each model across five different test sets for all cancers together and separate cancers. Percentages indicate the prevalence of each cancer type across the entire data set. The error bars indicate the minimum and maximum recall for each model/cancer across the five test sets.

### Results by Cancer Type

Across the population, different cancers occur at different rates and have varying propensities for metastasis^22^. Our dataset categorizes cancers based on their primary sites of origin. To evaluate the model’s performance in identifying metastasis across different cancer types, we stratified weighted precision, recall, and F1 score by cancer type (Table 3). We present direct comparison between the task-specific and the general LLM for each cancer type in Fig. 3, showing that the task-specific HiSAN model has superior performance across all cancer types.

Our evaluation reveals that the pathology reports associated with melanoma consistently achieve the highest predictive scores, ranging from an average F1 score of 0.863 for Llama3 to 0.878 for the HiSAN without the confidence threshold, to 0.964 for the HiSAN model with a threshold of 0.955. On the other hand, the ovarian/fallopian cancers present the most challenging reports to classify. Llama3 achieves a mean F1 score of 0.694, while HiSAN without thresholding achieves 0.733. The precision, recall, and F1 scores for ovarian/fallopian cancer show significant improvement when an abstention threshold is applied—increasing from 0.709 (precision), 0.763 (recall), and 0.733 (F1) to 0.919 (precision), 0.947 (recall), and 0.932 (F1). At a mean abstention rate of 0.578, we are able to identify and omit 92% of misclassified reports.

### Analysis of Knowledge Transfer Across Cancer Types

To determine whether our HiSAN model benefits more from being trained on multiple cancer types simultaneously (Figure 4A) or when dedicated models tailored to individual cancers are used, we trained separate HiSAN models for each cancer type (Figure 4B). This approach allows us to assess the trade-offs between leveraging shared knowledge across cancer types and specializing in the unique characteristics of each cancer type. Our analysis reveals that HiSAN models trained exclusively on individual cancer types generally underperform compared to those trained on the entire dataset when evaluated on the same cancer type (Table 2). This discrepancy is particularly pronounced for minority classes, such as ovarian/fallopian cancers (mean F1 improving from 0.577 to 0.709) and the heterogeneous “Other” category (mean F1 improving from 0.644 to 0.823), which benefit substantially from the shared model’s ability to transfer knowledge across cancer types.

**Figure 4:**
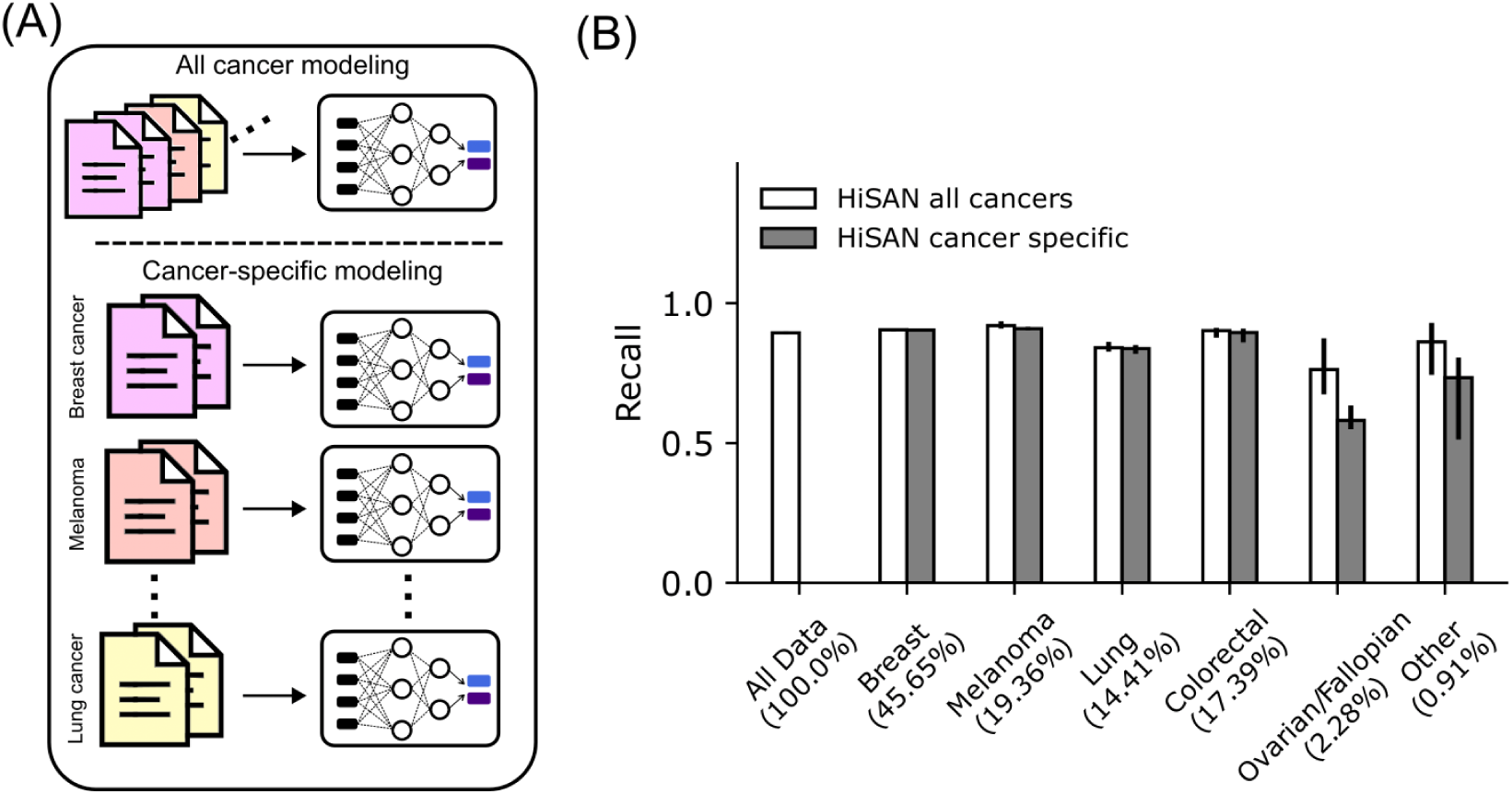
Modeling all cancers improves model performance compared to modeling each independently. (A) Top: all cancers are modeled by a single task-specific HiSAN model. Bottom: Each model trained only on the specific cancer type. (B) Mean recall of a model trained on all cancers simultaneously (white bars) and separately (dark bars) across five different test sets for all cancers together and separate cancers. Percentages indicate the prevalence of each cancer type across the entire data set. The error bars indicate the minimum and maximum recall for each model/cancer across the five test sets.

### Analysis and Review of the Undetermined Class

Another means of assessing our model is analyzing its confidence on the 2,226 *metastasis undetermined* reports. The HiSAN model abstains from making predictions on around 69% of these reports (see Table 4), effectively managing uncertain cases by deferring them for human review. The model shows high confidence with *metastasis negative* reports, flagging only 18% for review. However, for *metastasis positive* reports, it abstains on 48%, likely due to their complexity and fewer representative examples in the training data, reducing prediction confidence.

**Table 4.** Abstention rates for each class based on the 5-fold cross-validation.

| ***Threshold*** | ***Abstention Rate***<br>***(AR)***<br>*Mean (Low, High)* | ***Metastasis***<br>***Positive AR***<br>*Mean (Low, High)* | ***Metastasis***<br>***Negative AR***<br>*Mean (Low, High)* | ***Metastasis***<br>***Undetermined AR***<br>*Mean (Low, High)* |
| --- | --- | --- | --- | --- |
| 0.955 | 0.271 (0.232, 0.320) | 0.482 (0.410, 0.549) | 0.179 (0.156, 0.222) | 0.692 (0.498, 0.716) |

Of the 31% (N=690) of *metastasis undetermined* reports that the model classified as *metastasis positive* or *metastasis negative* with high confidence, we conducted a secondary review on a random sample of 66. Among these, 21 reports should have been abstained on due to insufficient information for a definitive determination. For the remaining 45 reports, our model achieved 97.8% accuracy compared to the final review labels, with a recall of 0.97 for *metastasis negative* and 1.0 for *metastasis positive*. These results highlight the model’s strength in resolving uncertain cases, offering valuable insights where human reviewers had initially expressed uncertainty.

## Discussion

This study presents a robust deep-learning approach for the automated extraction of metastasis information from unstructured pathology reports. Our work provides three key insights into the potential of deep learning for population-level metastasis identification.

First, our study is the first to integrate multiple cancer types from many pathology laboratories into a single deep learning algorithm for metastasis identification. This diversity introduces challenges; specifically, pathology reports from different laboratories vary significantly in stylistic and structural formatting. However, the HiSAN model learns to overcome these differences and only uses the relevant information in the reports. Furthermore, by including multiple cancer types, our model has an opportunity to learn a rich and nuanced representation of vocabulary associated with metastasis. We hypothesize that this improved performance is attributed to shared features common to all cancers, which aid in distinguishing between *metastasis negative* and *metastasis positive* cases. To validate this, we trained a HiSAN model on each cancer type alone and observed that it performed worse compared to when it was trained on the entire dataset (Table 2, Fig. 4). Ovarian/fallopian cancers, making up 2.28% of the data, benefit the most from knowledge transfer across cancer types (Fig. 4).

Second, our study demonstrates that a small, task-specific model can be better suited than a general-purpose large language model for metastasis identification. We found that the HiSAN performs 5 percentage points better than Llama3 in the three-class classification problem, raising concerns about the accuracy of general-purpose models utilized for medical applications^12,13^.

Furthermore, LLMs are trained on vast amounts of internet data, which can lead to unpredictable behavior and potential trust issues. Llama3 is resource-intensive, requiring significantly more computational power and memory compared to HiSAN. Despite its lower accuracy, Llama3 offers several advantages. Its versatility allows it to perform a wide range of tasks without requiring task-specific training examples, making it less vulnerable to issues like class imbalance and dataset biases. However, while these benefits are promising, we believe that the current accuracy of the LLM for zero-shot classification in medical contexts is not yet sufficient for practical deployment.

Third, our approach addresses the often-overlooked need for UQ in medical machine learning. We incorporate an abstention mechanism^23^, which serves as a crucial safeguard in real-world settings, particularly when patient data deviates from the patterns well-represented in the training set. The effectiveness of our method is underscored by its performance on ovarian and fallopian cancers before (Table 2) and after (Table 4) applying the abstention threshold. We were able to identify and omit 92% of misclassified reports, resulting in a notable increase of 0.201 in the F1 score, thereby enhancing the reliability of our model. Furthermore, we leverage data labeled as *undetermined* to assess high-confidence model predictions, recognizing that this category likely encompasses greater variability and functions as a catchall during manual labeling. Notably, upon secondary review, we discovered that our model demonstrated the capability to correct misclassifications originally made by human reviewers, underscoring its potential to serve as a decision-support tool in complex diagnostic scenarios.Together, these findings demonstrate the importance of tailored models and careful UQ in improving the reliability and accuracy of metastasis identification in large-scale clinical datasets.

In conclusion, we developed an accurate deep learning model for automatically identifying metastasis from pathology reports across diverse hospitals, independent pathology laboratories, geographical regions, and cancer types. We benchmarked our method against Llama3, demonstrating superior accuracy across all cancer types. We also found that our model correctly identifies ambiguous cases that warrant human review, enhancing its suitability for clinical applications.

## Methods

### Models

We utilize two modeling approaches: (1) a task-specific Hierarchical Self-Attention Network (HiSAN)^3^, which is a deep neural network trained from scratch for our specific classification task, and (2) the Llama3 pre-trained LLM, which conducts classification through prompting. We tailor the formulation of the classification problem to the strengths and weaknesses of these modeling approaches.

#### Task-Specific Approach

The HiSAN is a neural network architecture that incrementally constructs representations of a given report from the word level, through the sentence level, up to the document level using a self-attention mechanism. The final document-level representation is then fed into a linear classification layer, which predicts the probabilities of various document labels. This approach yields a model trained on a corpus of cancer patients’ pathology reports to identify metastasis occurring after the initial diagnosis. For this approach, we exclude from the training sets the potentially confusing reports that oncology data specialists labeled as *metastasis undetermined* (3.7% of all the reports). We frame the task as a binary classification problem focused on distinguishing between *metastasis positive* and *metastasis negative* conditions. Model training details and further justification for this reformulation are provided in the Supplementary Material.

#### General Model Approach

We utilize the pre-trained (i.e., trained on a large and diverse set of internet data that is not oncology-specific) Llama3 LLM. It is capable of understanding and generating human-like text and has been designed and trained to handle various NLP tasks through prompting techniques. We prompt the LLM to distinguish between the *metastasis positive, metastasis negative*, and *metastasis undetermined* categories and rely on its broader contextual understanding and reasoning capabilities.

For our classification task, the LLM is provided a prompt along with a pathology report. The model analyzes the report and generates the required output based on the instructions in the prompt. We test two prompts to optimize Llama3 performance: Prompt 1 mimics the instructions provided to human annotators, whereas Prompt 2 (see Supplementary Material) employs a technique in which the LLM is asked to take the role of an expert^19^. We found that Prompt 1 results in higher F1 scores on validation sets and thus use it in our final evaluation.

### Prompt 1

*Classify the provided pathology reports. Return 1 for one or more metastasis, return 0 for no metastasis reported on this path, return 8 for undetermined whether metastasis or subsequent primary or direct invasion from known primary. Provide output in the format: Answer: 0,1,8*

### Data Preprocessing

Data for our experiments was prepared using BARDI, an AI-readiness package for clinical text preprocessing^24^ (Figure 1: Data Preprocessing) The pathology reports were first normalized through a series of custom regular expressions and then split into word-based tokens. We employ 5-fold cross-validation to provide more accurate estimates of our models’ performance. In contrast to an entirely random split, we ensure that pathology records associated with the same patient are entirely contained within a single fold.

### Model Training

The Hierarchical Self-Attention Network (HiSAN) model was trained on 80% of the pathology reports that were labeled as either indicative of metastasis or not for all cancer types at all registries. The model was trained using FrESCO^25^ with a learning rate of 10^-4^, patience of 5, and a dropout rate of 0.1, and it converged after 15–20 epochs (Figure 1: Model Training). For the HiSAN architecture, which consisted of 8 attention heads, we selected *max words per line* to be 15, *attention dimension per head* to be 50, and *max document length* to be 3,000 words.

### Uncertainty Quantification

Uncertainty quantification (UQ) is essential for decision-making with classification models. In the task-specific approach, prediction confidence is indicated by the softmax score, representing the probability of a report belonging to a specific class (Figure 1: Uncertainty Quantification). By setting a confidence threshold, reports likely to be correct can be retained, while uncertain ones are discarded. This study explores the trade-offs between performance metrics and the abstention rate, where flagged reports are reviewed by humans. Minimizing the number of reports needing manual review is a crucial goal, given the time-consuming nature of the process. Because research investigating UQ in the context of LLMs is still in its early stages, we have not explored it in the present study (see Supplementary Material).

### Threshold selection

Because we care about accurately identifying metastasis, we aim to determine a confidence threshold *α*. Data samples *x* with a maximum softmax score *max* (*f*_θ_(*x_i_*)) > α are to be automatically classified, and those below the threshold deferred for manual review. Therefore, we aim to maximize the recall of the metastasis positive cases while minimizing the number of reports that need to be manually reviewed. We observed that the recall-abstention curve has a characteristic “knee” shape, in which further increases in the threshold result in a relatively minimal increases in recall. The output of the HiSAN *f*_θ_(*x_i_*) gives the probability of each class.

The recall for metastasis positive is

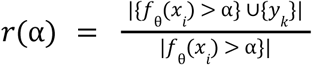

where {*y_k_*} is the set of labels, {*f*_θ_(*x_i_*) > α} is the set of pathology reports predicted to be metastasis with confidence more than α. The knee-finding algorithm outlined in^21^ determines the knee point of *r*(α), α by identifying the point of maximum curvature. This is estimated by

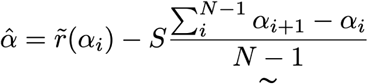

where *S* is the sensitivity parameter which we set to 1. *r_i_*(α) are the local maxima determined by the kneedle algorithm.

## Data Availability

The data used in this study consists of patient health information, which is subject to strict privacy and confidentiality regulations. Due to the sensitive nature of the data and compliance with applicable data protection laws, it is not publicly available. Access to this data is restricted to authorized researchers under institutional review board (IRB) approval and appropriate data use agreements. As a result, it cannot be shared or used outside the context of the approved study.

## Acknowledgements

We would like to thank Dr. Ardy Davarifar for his insightful feedback and constructive suggestions on the manuscript.

This work has been supported in part by the US Department of Energy (DOE) and the NCI of the National Institutes of Health. This work was performed under the auspices of the DOE by Oak Ridge National Laboratory under Contract DE-AC05-00OR22725.

## Supplementary Material

### Data Preprocessing

Data for our experiments was prepared using BARDI, an AI-readiness package for clinical text preprocessing^24^. The pathology reports were first normalized through a series of custom regular expressions and then split into word-based tokens. We employ 5-fold cross-validation to provide more accurate estimates of our models’ performance. In contrast to an entirely random split, we ensure that pathology records associated with the same patient are entirely contained within a single fold.

### Model Training

The Hierarchical Self-Attention Network (HiSAN) model was trained on 80% of the pathology reports that were labeled as either indicative of metastasis or not for all cancer types at all registries. The model was trained using FrESCO^25^ with a learning rate of 10^-4^, patience of 5, and a dropout rate of 0.1, and it converged after 15–20 epochs. For the HiSAN architecture, which consisted of 8 attention heads, we selected *max words per line* to be 15, *attention dimension per head* to be 50, and *max document length* to be 3,000 words.

### Justification for Reformulating the Classification into Two–Class Problem

Learning a rich representation of the *metastasis undetermined* reports is challenging due to the ambiguity of the concept and the limited number of available samples in the dataset. By training the model without these reports, we aim to ensure that it can learn to effectively discern features indicative of these two conditions with high confidence. After training, we anticipate that the model will exhibit uncertainty when faced with challenging and ambiguous reports, such as those in the *metastasis undetermined* category. For that reason, we introduce an abstention mechanism that allows the model to refrain from making predictions when its confidence is low. This abstention approach flags reports from the *undetermined* class for manual review. More broadly, it allows our model to reduce the incidence of false positives and false negatives by choosing not to classify ambiguous cases.

### Large Language Model Prompting

To run the Llama3 large language model (LLM), we utilized HuggingFace’s *pipeline* module with sampling turned off for deterministic outputs. We used two prompts.

#### Prompt 1

*Classify the provided pathology reports. Return 1 for one or more metastasis, return 0 for no metastasis reported on this path, return 8 for undetermined whether metastasis or subsequent primary or direct invasion from known primary. Provide output in the format: Answer: 0, 1, 8*.

#### Prompt 2

*You are a specialist tasked with annotating pathology reports at a SEER registry. Your role involves reading and understanding pathology reports. You look for key terms and phrases related to metastasis and classify each report as indicating metastasis, no metastasis, or inconclusive. Return 1 for metastasis, 0 for no metastasis, 8 for inconclusive. Provide output in the format: Answer: 0, 1, or 8*.

We find that Prompt 1 outperforms Prompt 2. The results for the Prompt 2 show an F1 score ranging from 0.775 to 0.802, with mean of 0.790.

### Privacy Considerations When Using LLMs for Clinical Applications

Although numerous models have been trained for medical applications, the interfaces likely to be used by a clinician day-to-day are those that enable easy interaction (e.g., OpenAI’s ChatGPT).

However, exposing private patient data to companies such as OpenAI is illegal and unethical. Comparable open-source models such as Llama enable researchers to understand how accurate a system like ChatGPT *would* be while running the model in a safe and secure environment.

### On Uncertainty Quantification for LLMs

Research into uncertainty quantification for LLMs is still in its early stages. Some of the recently proposed approaches rely on the nondeterministic nature of LLMs, in which the next token prediction can be sampled across more or less likely choices using a parameter called *temperature*. While performing multiple inference calls on the same case, we can quantify the confidence via voting. These methods complicate the automation of using LLMs because structured output is required. At high temperatures, the outputs are less likely to follow the prescribed structures. Additionally, repeated runs for the same prompt increase computational expense, making it impractical for high-throughput applications.

**Table S1:** Counts of Metastasis Negative, Metastasis Positive, and Undetermined labeled reports in each fold. Employing 5-fold cross-validation, we train five distinct models on 4 of 5 folds, with the 5th designated for validation and testing. The folds are created with patient-level stratification.

|  | *Training Set*<br>(Metastasis Negative /<br>Metastasis Positive /<br>Undetermined) | *Validation Set*<br>(Metastasis Negative /<br>Metastasis Positive /<br>Undetermined) | *Test Set*<br>(Metastasis Negative /<br>Metastasis Positive /<br>Undetermined) |
| --- | --- | --- | --- |
| **Experiment 1** | 48,396<br>(34,695 / 11,882 / 1,819) | 6,137<br>(4,346 / 1,578 / 213) | 5,938<br>(4,245 / 1,499 / 194) |
| **Experiment 2** | 48,456<br>(34,614 / 12,074 / 1,768) | 6,089<br>(4,462 / 1,419 / 208) | 5,926<br>(4,210 / 1,466 / 250) |
| **Experiment 3** | 48,415<br>(34,690 / 11,943 / 1,782) | 6,081<br>(4,341 / 1,519 / 221) | 5,975<br>(4,255 / 1,497 / 223) |
| **Experiment 4** | 48,167<br>(34,566 / 11,856 / 1,745) | 6,087<br>(4,374 / 1,575 / 268) | 6,217<br>(4,374 / 1,575 / 213) |
| **Experiment 5** | 48,550<br>(34,561 / 12,081 / 1,790) | 6,131<br>(4,432 / 1,482 / 217) | 5,890<br>(4,275 / 1,396 / 219) |

**Table S2:** Results for HiSAN on two-class classification (metastasis negative/ metastasis positive).

|  | ***HiSAN Evaluation on Two Classes*** |  |  |
| --- | --- | --- | --- |
|  | ***Precision***<br>*Mean*<br>*Low, High* | ***Recall***<br>*Mean*<br>*Low, High* | ***F1***<br>*Mean*<br>*Low, High* |
| ***All Data*** | 0.932<br>0.924, 0.938 | 0.930<br>0.920, 0.938 | 0.931<br>0.921, 0.938 |
| Breast | 0.941<br>0.933, 0.946 | 0.937<br>0.925, 0.943 | 0.938<br>0.926, 0.944 |
| Melanoma | 0.947<br>0.939, 0.959 | 0.944<br>0.937, 0.956 | 0.945<br>0.938, 0.957 |
| Lung | 0.905<br>0.889, 0.920 | 0.904<br>0.889, 0.921 | 0.904<br>0.889, 0.920 |
| Colorectal | 0.932<br>0.909, 0.945 | 0.931<br>0.908, 0.944 | 0.931<br>0.908, 0.944 |
| Ovarian/<br>Fallopian | 0.827<br>0.744, 0.920 | 0.824<br>0.744, 0.919 | 0.824<br>0.744, 0.919 |
| Other | 0.921<br>0.879, 0.955 | 0.914<br>0.879, 0.952 | 0.916<br>0.879, 0.950 |

## References

1. Cancer Statistics - NCI. April 2, 2015. Accessed July 22, 2024. https://www.cancer.gov/about-cancer/understanding/statistics

2. Chandrashekar M, Lyngaas I, Hanson HA, Gao S, Wu XC, Gounley J. Path-BigBird: An AI-Driven Transformer Approach to Classification of Cancer Pathology Reports. JCO Clin Cancer Inform. 2024;(8):e2300148. doi:10.1200/CCI.23.00148

3. Gao S, Qiu JX, Alawad M, et al. Classifying cancer pathology reports with hierarchical self-attention networks. Artif Intell Med. 2019;101:101726. doi:10.1016/j.artmed.2019.101726

4. Lee J, Yoon W, Kim S, et al. BioBERT: a pre-trained biomedical language representation model for biomedical text mining. Bioinformatics. 2020;36(4):1234–1240. doi:10.1093/bioinformatics/btz682

5. Yang X, Chen A, Pour Nejatian N, et al. A large language model for electronic health records. Npj Digit Med. 2022;5(1):1–9. doi:10.1038/s41746-022-00742-2

6. Li Y, Wehbe RM, Ahmad FS, Wang H, Luo Y. Clinical-Longformer and Clinical-BigBird: Transformers for long clinical sequences. Published online April 15, 2022. doi:10.48550/arXiv.2201.11838

7. Soysal E, Warner JL, Denny JC, Xu H. Identifying Metastases-related Information from Pathology Reports of Lung Cancer Patients. AMIA Summits Transl Sci Proc. 2017;2017:268–277.

8. Batch KE, Yue J, Darcovich A, et al. Developing a Cancer Digital Twin: Supervised Metastases Detection From Consecutive Structured Radiology Reports. Front Artif Intell. 2022;5. doi:10.3389/frai.2022.826402

9. Banerjee I, Bozkurt S, Caswell-Jin JL, Kurian AW, Rubin DL. Natural Language Processing Approaches to Detect the Timeline of Metastatic Recurrence of Breast Cancer. JCO Clin Cancer Inform. 2019;(3):1–12. doi:10.1200/CCI.19.00034

10. Groot OQ, Bongers MER, Karhade AV, et al. Natural language processing for automated quantification of bone metastases reported in free-text bone scintigraphy reports. Acta Oncol. 2020;59(12):1455–1460. doi:10.1080/0284186X.2020.1819563

11. Sangariyavanich E, Ponthongmak W, Tansawet A, et al. Systematic review of natural language processing for recurrent cancer detection from electronic medical records. Inform Med Unlocked. 2023;41:101326. doi:10.1016/j.imu.2023.101326

12. Sushil M, Zack T, Mandair D, et al. A comparative study of large language model-based zero-shot inference and task-specific supervised classification of breast cancer pathology reports. J Am Med Inform Assoc. Published online June 20, 2024:ocae146. doi:10.1093/jamia/ocae146

13. Williams CYK, Zack T, Miao BY, et al. Use of a Large Language Model to Assess Clinical Acuity of Adults in the Emergency Department. JAMA Netw Open. 2024;7(5):e248895. doi:10.1001/jamanetworkopen.2024.8895

14. Shah NH, Entwistle D, Pfeffer MA. Creation and Adoption of Large Language Models in Medicine. JAMA. 2023;330(9):866–869. doi:10.1001/jama.2023.14217

15. Perlis RH, Fihn SD. Evaluating the Application of Large Language Models in Clinical Research Contexts. JAMA Netw Open. 2023;6(10):e2335924. doi:10.1001/jamanetworkopen.2023.35924

16. Longwell JB, Hirsch I, Binder F, et al. Performance of Large Language Models on Medical Oncology Examination Questions. JAMA Netw Open. 2024;7(6):e2417641. doi:10.1001/jamanetworkopen.2024.17641

17. Seoni S, Jahmunah V, Salvi M, Barua PD, Molinari F, Acharya UR. Application of uncertainty quantification to artificial intelligence in healthcare: A review of last decade (2013-2023). Comput Biol Med. 2023;165:107441. doi:10.1016/j.compbiomed.2023.107441

18. Zhang X, Talukdar N, Vemulapalli S, et al. Comparison of Prompt Engineering and Fine-Tuning Strategies in Large Language Models in the Classification of Clinical Notes. Published online February 8, 2024:2024.02.07.24302444. doi:10.1101/2024.02.07.24302444

19. Meskó B. Prompt Engineering as an Important Emerging Skill for Medical Professionals: Tutorial. J Med Internet Res. 2023;25:e50638. doi:10.2196/50638

20. Risk Factors: Age - NCI. April 29, 2015. Accessed July 22, 2024. https://www.cancer.gov/about-cancer/causes-prevention/risk/age

21. Satopaa V, Albrecht J, Irwin D, Raghavan B. Finding a “Kneedle” in a Haystack: Detecting Knee Points in System Behavior. In: 2011 31st International Conference on Distributed Computing Systems Workshops.; 2011:166–171. doi:10.1109/ICDCSW.2011.20

22. Budczies J, von Winterfeld M, Klauschen F, et al. The landscape of metastatic progression patterns across major human cancers. Oncotarget. 2014;6(1):570–583.

23. Kompa B, Snoek J, Beam AL. Second opinion needed: communicating uncertainty in medical machine learning. Npj Digit Med. 2021;4(1):1–6. doi:10.1038/s41746-020-00367-3

24. Krawczuk, Patrycja, Murdock, Dakota. BARDI: Batch-processing Abstraction for Raw Data Integration. Published online March 2024. doi:10.11578/dc.20240328.2

25. Spannaus A, Gounley J, Shekar MC, et al. FrESCO: Framework for Exploring Scalable Computational Oncology. J Open Source Softw. 2023;8(89):5345. doi:10.21105/joss.05345

